# A novel workflow for the safe and effective integration of AI as supporting reader in double reading breast cancer screening: A large-scale retrospective evaluation

**DOI:** 10.1101/2022.06.22.22276751

**Authors:** Annie Y. Ng, Ben Glocker, Cary Oberije, Georgia Fox, Jonathan Nash, Edith Karpati, Sarah Kerruish, Peter D. Kecskemethy

**Affiliations:** Kheiron Medical Technologies, London, United Kingdom; Department of Computing, Imperial College London, London, United Kingdom

## Abstract

**Objectives:** To evaluate the effectiveness of a novel strategy for using AI as a supporting reader for the detection of breast cancer in mammography-based double reading screening practice. Instead of replacing a human reader, here AI serves as the second reader only if it agrees with the recall/no-recall decision of the first human reader. Otherwise, a second human reader makes an assessment, enacting standard human double reading.

**Design:** Retrospective large-scale, multi-site, multi-device, evaluation study.

**Participants:** 280,594 cases from 180,542 female participants who were screened for breast cancer with digital mammography between 2009 and 2019 at seven screening sites in two countries (UK and Hungary).

**Main outcome measures:** Primary outcome measures were cancer detection rate, recall rate, sensitivity, specificity, and positive predictive value. Secondary outcome was reduction in workload measured as arbitration rate and number of cases requiring second human reading.

**Results:** The novel workflow was found to be superior or non-inferior on all screening metrics, almost halving arbitration and reducing the number of cases requiring second human reading by up to 87.50% compared to human double reading.

**Conclusions:** AI as a supporting reader adds a safety net in case of AI discordance compared to alternative workflows where AI replaces the second human reader. In the simulation using large-scale historical data, the proposed workflow retains screening performance of the standard of care of human double reading while drastically reducing the workload. Further research should study the impact of the change in case mix for the second human reader as they would only assess cases where the AI and first human reader disagree.

## Introduction

The implementation of AI as an independent reader in mammography-based double reading breast cancer screening has the potential to reduce workload while preserving and possibly improving accuracy for cancer detection as suggested in recent large-scale retrospective studies [1–6]. To further assess the effectiveness of AI in screening practice, current evidence needs to be complemented by investigating the impact of clinical deployment of AI [7]. Here it is important to evaluate different strategies for the integration of AI with the goal to optimise the interaction between AI and human readers, maximising the combined benefit while ensuring patient safety and minimising clinical and operational risks [8].

Various AI workflows in screening have been discussed in the literature, including AI replacing one or all human readers [3–5,9], AI for triaging and prioritisation (before human reading) [10–12], or AI as an extra reader for identifying cancers missed by human reading [10,13]. These different roles for AI in the screening pathway have profound implications on the amount of automation, associated risks, participants’ acceptance, regulatory approval, and the downstream effects of any human-AI interaction [14]. While AI used for triaging and decision-referral showed potential for drastically reducing workload [15,16], the fact that a large number of cases would not be assessed by any human reader poses clinical risks and may hinder its acceptance by screening participants [17]. A large majority of screening participants seem to agree that some level of human oversight is desired [18]. In this context, using AI as a standalone reader entirely replacing human readers seems unlikely to become a viable strategy for deployment in the foreseeable future [19].

AI serving as an independent second reader in a double reading setting appears to strike a good balance through its potential to reduce workload while preserving double reader accuracy, keeping the assurances of having at least one human expert reader to assess every individual case at all times. A recent systematic review, however, has remarked that current AI systems, when serving as an independent second reader, may increase arbitration rates which would have clinical and operational implications [14]. Here, we propose and evaluate a novel strategy for adopting AI as a supporting reader within a human double reading pathway. In this new workflow, the AI serves as the second reader if the AI prediction agrees with the first human reader’s opinion. Otherwise, the AI prediction is disregarded, and a second human reader is making an assessment, from which point the standard human double reading is enacted. This added safety net reduces the risks of using AI while retaining the potential to significantly reduce workload. Evaluating the effectiveness of this new workflow and comparing it to both the standard of care of human double reading and AI serving as an independent second reader is the objective of this simulation analysis.

## Methods

### Study design

The study is a retrospective evaluation using data from a recent large-scale clinical study [6]. The study data was used to simulate double reading performance using AI as an independent reader for the detection of breast cancer in full-field digital mammography (FFDM) images. A novel workflow of using AI as supporting reader (AI-SR) was simulated and compared to the historical human double reading (HDR) and AI serving as an independent second reader (AI-IR) (see Figure 1). AI-SR is a variation of the previously explored AI-IR workflow [3,6], specifically designed to avoid an increase in arbitration rates. All performance comparisons were determined on the same unenriched screening cohorts representative of data the AI would see in real-world deployments. The original study protocol detailing the original inclusion/exclusion criteria and target performance metrics was established prior to opening the original study, which is presented elsewhere [6]. In the present evaluation, cases with a history of breast cancer (1.7%) were included to assess performance on a wider range of patients.

**Figure 1:**
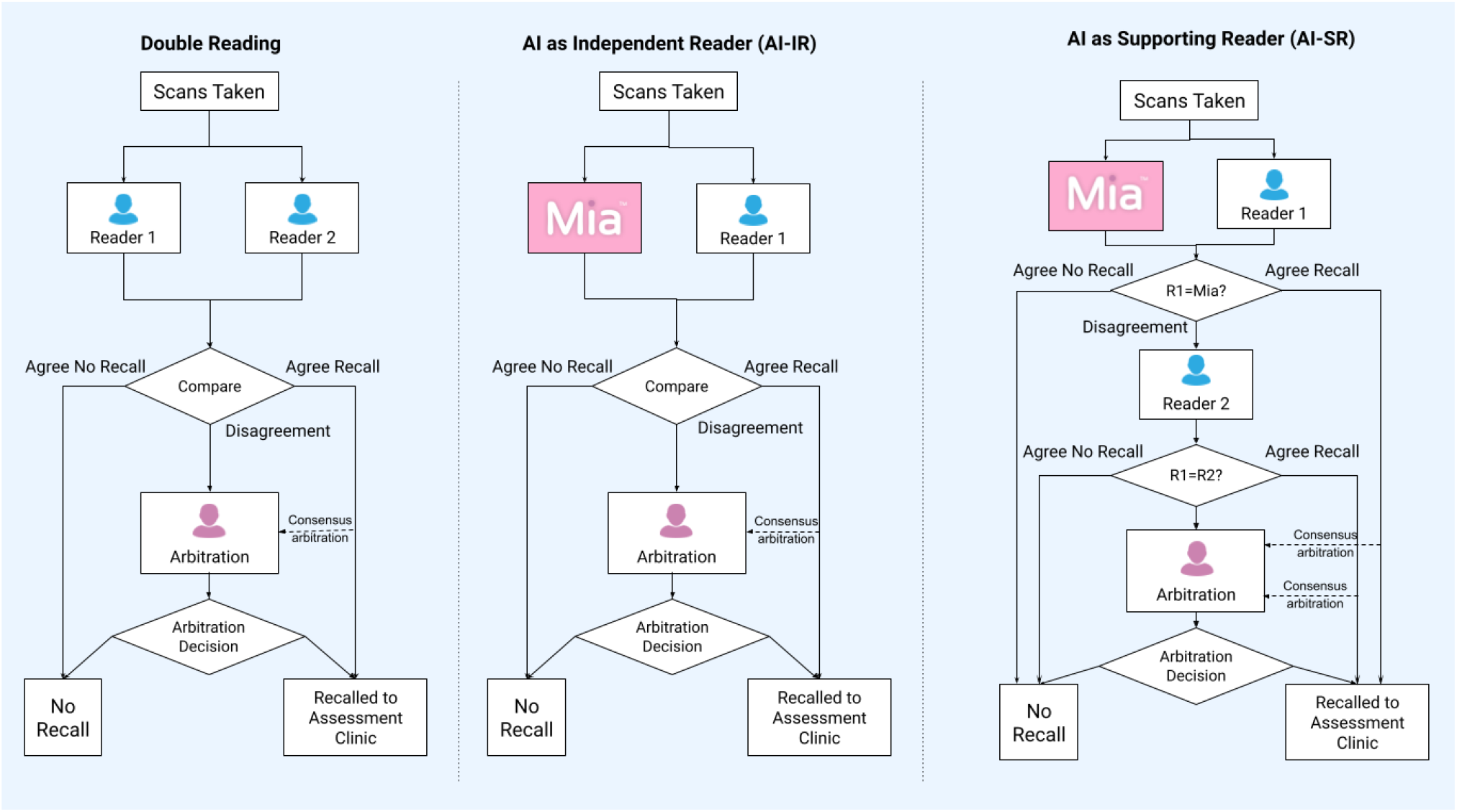
Diagrams illustrating the three workflows compared in this study

### Study population

The study population consisted of 280,594 cases from 180,542 female participants who were invited to breast cancer screening between 2009 and 2019. The sample was representative of the population to which the AI system would be applied to in real-world screening practice. De-identified cases were collected from seven sites in the United Kingdom and Hungary. The three UK centres included Leeds Teaching Hospital NHS Trust (LTHT), Nottingham University Hospitals NHS Trust (NUH), and United Lincolnshire Hospitals NHS Trust (ULH). All three centres participate in the UK NHS Breast Cancer Screening Programme and adhere to a three-year screening interval, with women between 50 and 70 years old invited to participate. A small cohort of women between 47 and 49 years, and 71 and 73 years old who were eligible for the UK age extension trial (Age X) were also included [20]. The Hungarian centre, MaMMa Klinika (MK), involved four sites, Budapest (KAP), Kecskemét (KKM), Szekszárd (SZE), Szolnok (SZO), and corresponding mobile screening units (BUS), which follow a two-year screening interval and invite women aged 45 to 65. Across all sites, women outside the regional screening programme age range, who chose to participate as per standard of care (opportunistic screening) were also included. Screening cases were acquired from the dominant mammography hardware vendor at each site: Hologic (at LTHT), GE Healthcare (NUH), Siemens (ULH), and IMS Giotto (MK).

Positive cases were pathology-proven malignancies confirmed by fine needle aspiration cytology (FNAC), core needle biopsy (CNB), vacuum-assisted core biopsy (VACB) and/or histology of the surgical specimen. All negatives had evidence of a three-year negative follow-up result. Further details on ground truthing, including subsample definitions are reported elsewhere [6].

### AI system

The AI system employed in this study (Mia™ version 2.0.1, Kheiron Medical Technologies) was previously assessed in a large-scale retrospective study [6]. The AI system works with standard DICOM (Digital Imaging and Communications in Medicine) cases as inputs, analyses four images with two standard FFDM (craniocaudal and mediolateral oblique) views per breast. The output of the AI is a single binary recommendation per case of “recall” (for further assessment due to suspected malignancy) or “no recall” (until the next screening interval). The AI software version and its operating points were fixed prior to the study. None of the study data was used in any aspect of algorithm development.

### Standard of care human double reading

At all screening sites, the second human reader had access, at their discretion, to the opinion of the first human reader. In cases of disagreement, an arbitration, performed by a single or group of radiologists, made the final decision. When the opinions of the first and second human reader agreed “no recall”, a definitive “no recall” decision was reached. When the opinions agreed “recall”, a “recall” decision was reached, or an arbitration performed by a single or group of radiologists made the definitive “recall” or “no recall” decision, depending on the site’s local practice.

### Double reading with an AI system

Previous studies [3,6] considered fully replacing the second human reader with AI as an independent reader (AI-IR) which was simulated by combining the (historical) first human reader’s opinion with the AI’s prediction. This workflow has direct implications on the arbitration process, as in the case of disagreement, human arbitrators would need to consider a human reader’s opinion together with an AI prediction for making a final decision instead of relying on opinions from two human readers as available in the standard of care human double reading (HDR).

In the proposed workflow of using AI as a supporting reader (AI-SR), the interaction between AI and human assessment is limited. Here, the first human reader’s opinion is the final “recall” or “no-recall” decision if it agrees with the AI’s prediction. In case of disagreement, the AI’s prediction is disregarded, and a second human reader makes an assessment. Thus, only human reader opinions are considered whenever arbitration is necessary (which is the case when the two human readers disagree). Similar to the AI-IR workflow, the AI-SR is simulated using the historical first and human reader’s opinion in this evaluation. Figure 1 illustrates the different workflows of HDR, AI-IR, and AI-SR compared in this study.

### Statistical analysis

Performance of historical HDR and the simulated use of AI was measured in terms of recall rate (RR), cancer detection rate (CDR), sensitivity, specificity, and positive predictive value (PPV). For these metrics, bootstrapping was used to calculate 95% confidence intervals. Non-inferiority was defined to rule out a relative difference of more than 10% in the direction of reduced performance with a 97.5% confidence and an alpha of 2.5%. Superiority was tested when noninferiority was passed and was also based on the same confidence intervals and alpha. Operational performance in terms of workload reduction was assessed as arbitration rate (the rate of disagreement between the first and second readers) and number of cases requiring second human reading.

## Results

### Study population characteristics

Table 1 presents characteristics of the study population. Of the 280,594 total cases, there were 2783 (0.99%) positives overall (historically detected), with 2397 (0.85%) screen-detected positives (in-line with screening expectations) and 386 (0.18%) interval cancers (ICs). From those, 293 (0.10%) were three-year ICs for the UK sample, and 93 (0.03%) were two-year ICs for the Hungarian sample. A breakdown per clinical site is provided in the supplementary material (see Table S1).

**Table 1:** Population characteristics

| Variable |  | Number of cases | Proportion of study population |
| --- | --- | --- | --- |
|  | Total | 280,594 | 100.0% |
| Country | UK | 194,145 | 69.2% |
|  | HU | 86,449 | 30.8% |
| Center / Vendor | NUH / GE (UK) | 69,621 | 24.8% |
|  | LTHT / Hologic (UK) | 65,098 | 23.2% |
|  | ULH / SIEMENS (UK) | 59,426 | 21.2% |
|  | KAP / IMS Giotto (HU) | 27,577 | 9.8% |
|  | BUS / IMS Giotto (HU) | 23,901 | 8.5% |
|  | SZO / IMS Giotto (HU) | 19,266 | 6.9% |
|  | KKM / IMS Giotto (HU) | 11,721 | 4.2% |
|  | SZE / IMS Giotto (HU) | 3,984 | 1.4% |
| Age (Year) | < 40 | 485 | 0.2% |
|  | 40 - 49 | 37,822 | 13.5% |
|  | 50 - 59 | 115,341 | 41.1% |
|  | 60 - 69 | 100,445 | 35.8% |
|  | 70 - 79 | 24,674 | 8.8% |
|  | 80 - 89 | 1,806 | 0.6% |
|  | 90+ | 21 | 0.01% |
| Positives | Total positives | 2,783 | 0.99% |
|  | Screen-detected positives total | 2,397 | 0.85% |
|  | Screen-detected positives UK | 1,718 | 0.61% |
|  | Screen-detected positives HU | 679 | 0.24% |
|  | Three-year ICs from UK* | 293 | 0.10% |
|  | Two-year ICs from HU* | 93 | 0.03% |

Abbreviations: HU = Hungary, UK = United Kingdom, KAP= Budapest, KKM = Kecskemét, LTHT = Leeds Teaching Hospital NHS Trust, NUH = Nottingham University Hospitals NHS Trust, SZE = Szekszárd, SZO = Szolnok, BUS = Szűrőbusz, ULH = United Lincolnshire Hospitals NHS Trust, IC = interval cancer
\*For calculation of sensitivity, screen-detected positives UK and three-year ICs from UK were used for UK sites and screen-detected positives HU and two-year ICs from HU were used for HU sites

### Cancer detection performance

Table 2 presents the average cancer detection performance separately for the UK and Hungarian sample in terms of RR, CDR, sensitivity, specificity, and PPV for the historical HDR and the simulated use of AI. Note, the cancer detection performance of the AI system in the simulation is the same for the AI-IR and AI-SR workflows.

**Table 2:** Cancer detection performance per country

| UK |  |  |  |  |  |  |  |  |
| --- | --- | --- | --- | --- | --- | --- | --- | --- |
|  | Historical HDR |  |  | Simulated AI-IR / AI-SR |  |  | Test AI vs HDR |  |
| Performance metric | Value | LBCI | UBCI | Value | LBCI | UBCI | Ratio | Difference |
| Recall rate | 3.84 | 3.75 | 3.92 | 3.82 | 3.73 | 3.91 | Non-inferior | Non-inferior |
| CDR | 8.93 | 8.52 | 9.35 | 8.71 | 8.31 | 9.12 | Non-inferior | Non-inferior |
| Sensitivity | 86.23 | 84.81 | 87.72 | 84.09 | 82.55 | 85.69 | Non-inferior | Non-inferior |
| Specificity | 96.98 | 96.83 | 97.13 | 97.05 | 96.91 | 97.19 | Superior | Superior |
| PPV | 23.28 | 22.39 | 24.19 | 22.82 | 21.92 | 23.70 | Non-inferior | Non-inferior |
| HU |  |  |  |  |  |  |  |  |
|  | Historical HDR |  |  | Simulated AI-IR / AI-SR |  |  | Test AI vs HDR |  |
| Performance metric | Value | LBCI | UBCI | Value | LBCI | UBCI | Ratio | Difference |
| Recall rate | 11.75 | 11.54 | 11.96 | 10.35 | 10.16 | 10.55 | Superior | Superior |
| CDR | 7.92 | 7.35 | 8.51 | 7.83 | 7.25 | 8.41 | Non-inferior | Non-inferior |
| Sensitivity | 88.73 | 86.33 | 90.88 | 87.69 | 85.25 | 89.85 | Non-inferior | Non-inferior |
| Specificity | 94.45 | 94.08 | 94.80 | 95.58 | 95.25 | 95.92 | Superior | Superior |
| PPV | 6.74 | 6.27 | 7.23 | 7.57 | 7.03 | 8.10 | Superior | Superior |

Abbreviations: UK = United Kingdom, HU = Hungary, HDR = human double reading, AI-IR = AI independent reader, AI-SR = AI supporting reader, LBCI = lower bound 95% confidence interval, UBCI = upper bound 95% confidence interval, CDR = cancer detection rate (per 1000 screens), PPV = positive predictive value

For the UK sample, the RR is 3.84% (95% CI 3.75 to 3.92) and CDR is 8.93% (8.52 to 9.35) for HDR, compared with a RR of 3.82% (3.73 to 3.91) and CDR of 8.71% (8.31 to 9.12) for AI-IR/SR. The sensitivity is 86.23% (84.81 to 87.72) with a specificity of 96.98% (96.83 to 97.13) for HDR, compared with a sensitivity of 84.09% (82.55 to 85.69) and specificity of 97.05% (96.91 to 97.19) for AI-IR/SR. The PPV is 23.28% (22.39 to 24.19) for HDR and 22.82% (21.92 to 23.70) for AI-IR/SR. The use of AI is non-inferior on RR, CDR, sensitivity, and PPV, and superior on specificity compared to HDR.

For the Hungarian sample, the RR is 11.75% (11.54 to 11.96) and CDR is 7.92% (7.35 to 8.51) for HDR, compared with a RR of 10.35% (10.16 to 10.55) and CDR of 7.83% (7.25 to 8.41) for AI-IR/SR. The sensitivity is 88.73% (86.33 to 90.88) with specificity of 94.45% (94.08 to 94.80) for HDR, compared with a sensitivity of 87.69% (85.25 to 89.85) and specificity of 95.58% (95.25 to 95.92) for AI-IR/SR. The PPV is 6.74% (6.27 to 7.23) for HDR and 7.57% (7.03 to 8.10) for AI-IR/SR. The use of AI is non-inferior on CDR and sensitivity, and superior on RR, specificity, and PPV.

A breakdown of the results per clinical site is provided in the supplementary material (see Table S2).

### Operational performance

For the standard of care HDR, all cases are read by a first and second human reader from which 9,655 (3.40%) cases were referred to arbitration due to disagreement between the human readers. For the simulated AI-IR workflow where the AI is fully replacing the second human reader there were 35,199 (12.50%) cases referred to arbitration due to disagreement between the first human reader and the AI prediction. For the proposed AI-SR workflow, these AI discordant cases would be referred to a second human reader, from which 5,056 (1.80%) cases were referred to arbitration due to disagreement between the human readers. When comparing HDR with AI-SR, the simulation shows a potential operational benefit for AI-SR with a reduction in arbitration rate of 47.63% and a reduction in the number of cases requiring second human reading of 87.50%. The results are visually presented in Figure 2.

**Figure 2:**
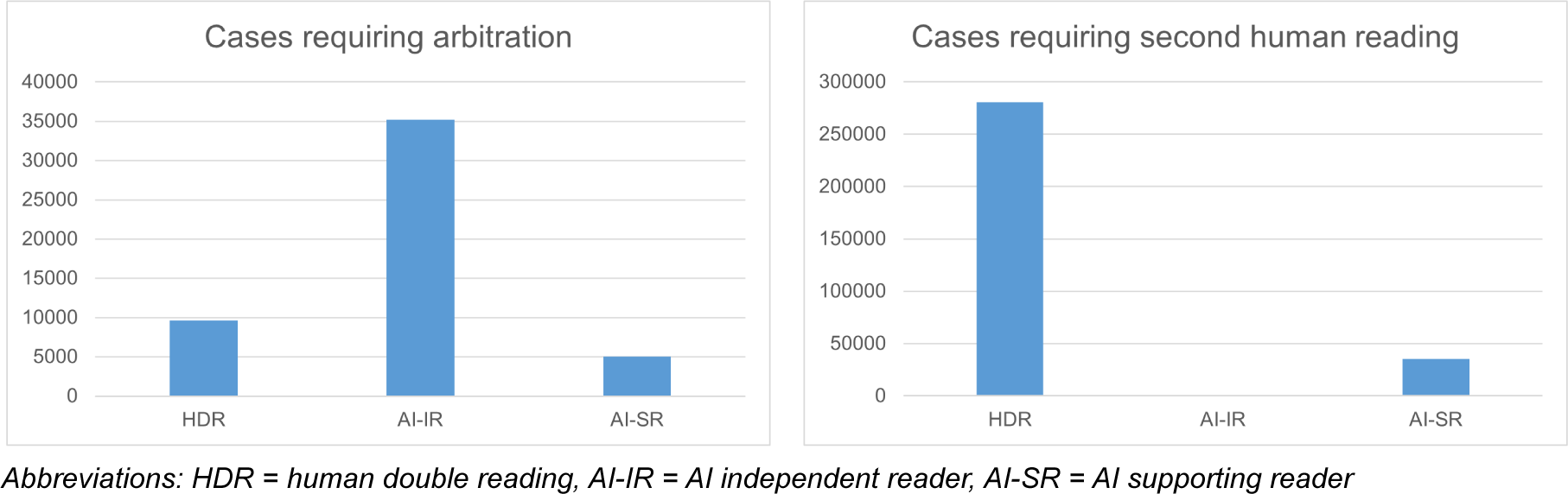
Workload in terms of number of cases for arbitration and second human reading

## Discussion

The proposed novel workflow of using AI as a supporting reader (AI-SR) retains cancer detection performance compared to the historical human double reading (HDR) in a simulation using a large-scale screening population. Compared to using AI as an independent reader (AI-IR) replacing the second human reader, the AI-SR workflow resulted in a significantly lower arbitration rate (12.50% vs 1.80%), which was also lower than the arbitration rate of HDR (3.40%). While AI-IR requires no second human reading among the cases eligible for AI processing, its increase in arbitration rate compared to HDR had been previously raised as a concern for the use of AI in screening [14]. Arbitration is a more time-consuming process than individual screening reads, and AI-IR comes with the implication of assessing a human reader’s opinion together with an AI prediction in the arbitration process. The AI prediction may be less interpretable and specific training for arbitrators may be required. The impact of this change in the arbitration process is unclear which has led to concerns regarding the clinical deployment of AI-IR [14]. Here, the proposed AI-SR workflow mitigates this concern as only human reader opinion’s would be considered during arbitration while the arbitration rate remains low and the number of cases requiring second human reading is still drastically reduced.

A main limitation of the study is that all results for the cancer detection and operational performance for the use of AI are based on a simulation using historical data. The simulation is exact in the case of AI-SR, while for AI-IR it is an approximation as the second human reader’s opinion was used when historical arbitration was not available, which was the case in 85.60% of arbitration cases. As we would expect that the second reader performance is worse than true arbitration, the approximation for AI-IR is likely to provide a lower bound of the real world performance. A further assumption is made that the historical second human reader behaviour is the same in HDR and AI-SR. However, as the second human reader in the AI-SR workflow would only assess cases where the first human reader and the AI disagree, there could be a change in the case mix as the AI discordant cases might be generally more difficult to assess which may impact the reader’s performance. Additional training may be required to adapt human readers to this change in the screening pathway.

A key strength of the study is the use of a large-scale, unenriched screening population with participants from two countries and multiple clinical sites including mobile units, and imaging data acquired on machines from four hardware vendors. This is important for the generalisability of the simulation results and their translation to screening practice.

Compared to AI-IR, one downside of the AI-SR workflow is that it does not provide the opportunity to identify more cancers that may have been missed by HDR. However, an important benefit is the added safety net for the clinical deployment by providing the assurance that the standard of care of human double reading is enacted whenever the first human reader and the AI prediction disagree. This may not only positively impact the participants’ acceptance for the integration of AI into screening practice, but may also help with gaining support for setting up prospective and randomised controlled trials involving AI systems. Prospective studies are an important next step to obtain further evidence about the benefit of AI in breast cancer screening. These studies will need to be carefully designed to ensure patient safety while minimising clinical and operational risks. Here, the proposed workflow of using AI as supporting reader could be a viable solution.

## Supporting information

STARD checklist

## Data Availability

The data for the current study are not publicly available. Due to reasonable privacy and ethical concerns, the imaging data cannot be distributed to researchers without ethical approval and research agreements with the original data providers.

## Supplementary Material

**Table S1:** Population characteristics per clinical site

| NUH (UK) |  |  |  |
| --- | --- | --- | --- |
| Variable |  | Number of cases | Proportion of study population |
|  | Total | 69621 | 100.00% |
| Age (Year) | < 40 | 0 | 0.00% |
|  | 40 - 49 | 6150 | 8.83% |
|  | 50 - 59 | 31156 | 44.75% |
|  | 60 - 69 | 25456 | 36.56% |
|  | 70 - 79 | 6473 | 9.30% |
|  | 80 - 89 | 386 | 0.55% |
|  | 90+ | 0 | 0.00% |
| Positives | Total positives | 734 | 1.05% |
|  | Screen-detected positives | 622 | 0.89% |
|  | Three-year ICs | 112 | 0.16% |
| LTHT (UK) |  |  |  |
| Variable |  | Number of cases | Proportion of study population |
|  | Total | 65098 | 100.00% |
| Age (Year) | < 40 | 0 | 0.00% |
|  | 40 - 49 | 6005 | 9.22% |
|  | 50 - 59 | 29961 | 46.02% |
|  | 60 - 69 | 23978 | 36.83% |
|  | 70 - 79 | 4994 | 7.67% |
|  | 80 - 89 | 160 | 0.25% |
|  | 90+ | 0 | 0.00% |
| Positives | Total positives | 622 | 0.96% |
|  | Screen-detected positives | 541 | 0.83% |
|  | Three-year ICs | 81 | 0.12% |
| ULH (UK) |  |  |  |
| Variable |  | Number of cases | Proportion of study population |
|  | Total | 59426 | 100.00% |
| Age (Year) | < 40 | 0 | 0.00% |
|  | 40 - 49 | 4059 | 6.83% |
|  | 50 - 59 | 24289 | 40.87% |
|  | 60 - 69 | 23636 | 39.77% |
|  | 70 - 79 | 6987 | 11.76% |
|  | 40 - 49 | 4059 | 6.83% |
|  | 80 - 89 | 455 | 0.77% |
|  | 90+ | 0 | 0.00% |
| Positives | Total positives | 655 | 1.10% |
|  | Screen-detected positives | 555 | 0.93% |
|  | Three-year ICs | 100 | 0.17% |
| **KAP (HU)** |  |  |  |
| **Variable** |  | **Number of cases** | **Proportion of study population** |
|  | Total | 27577 | 100.00% |
| Age (Year) | < 40 | 452 | 1.64% |
|  | 40 - 49 | 7360 | 26.69% |
|  | 50 - 59 | 8239 | 29.88% |
|  | 60 - 69 | 8390 | 30.42% |
|  | 70 - 79 | 2760 | 10.01% |
|  | 80 - 89 | 363 | 1.32% |
|  | 90+ | 13 | 0.05% |
| Positives | Total positives | 240 | 0.87% |
|  | Screen-detected positives | 195 | 0.71% |
|  | Two-year ICs | 45 | 0.16% |
| **BUS (HU)** |  |  |  |
| **Variable** |  | **Number of cases** | **Proportion of study population** |
|  | Total | 23901 | 100.00% |
| Age (Year) | < 40 | 3 | 0.01% |
|  | 40 - 49 | 6008 | 25.14% |
|  | 50 - 59 | 9659 | 40.41% |
|  | 60 - 69 | 7613 | 31.85% |
|  | 70 - 79 | 580 | 2.43% |
|  | 80 - 89 | 38 | 0.16% |
|  | 90+ | 0 | 0.00% |
| Positives | Total positives | 125 | 0.52% |
|  | Screen-detected positives | 111 | 0.46% |
|  | Two-year ICs | 14 | 0.06% |
| **SZO (HU)** |  |  |  |
| **Variable** |  | **Number of cases** | **Proportion of study population** |
|  | Total | 19266 | 100.00% |
| Age (Year) | < 40 | 29 | 0.15% |
|  | 40 - 49 | 4535 | 23.54% |
|  | 50 - 59 | 6664 | 34.59% |
|  | 60 - 69 | 6088 | 31.60% |
|  | 70 - 79 | 1672 | 8.68% |
|  | 80 - 89 | 272 | 1.41% |
|  | 90+ | 6 | 0.03% |
| Positives | Total positives | 283 | 1.47% |
|  | Screen-detected positives | 260 | 1.35% |
|  | Two-year ICs | 23 | 0.12% |

| KKM (HU) |  |  |  |
| --- | --- | --- | --- |
| Variable |  | Number of cases | Proportion of study population |
|  | Total | 11721 | 100.00% |
| Age (Year) | < 40 | 1 | 0.01% |
|  | 40 - 49 | 2720 | 23.21% |
|  | 50 - 59 | 4024 | 34.33% |
|  | 60 - 69 | 3895 | 33.23% |
|  | 70 - 79 | 969 | 8.27% |
|  | 80 - 89 | 111 | 0.95% |
|  | 90+ | 1 | 0.01% |
| Positives | Total positives | 97 | 0.83% |
|  | Screen-detected positives | 86 | 0.73% |
|  | Two-year ICs | 11 | 0.09% |
| SZE (HU) |  |  |  |
| Variable |  | Number of cases | Proportion of study population |
|  | Total | 3984 | 100.00% |
| Age (Year) | < 40 | 0 | 0.00% |
|  | 40 - 49 | 985 | 24.72% |
|  | 50 - 59 | 1349 | 33.86% |
|  | 60 - 69 | 1389 | 34.86% |
|  | 70 - 79 | 239 | 6.00% |
|  | 80 - 89 | 21 | 0.53% |
|  | 90+ | 1 | 0.03% |
| Positives | Total positives | 27 | 0.68% |
|  | Screen-detected positives | 27 | 0.68% |
|  | Two-year ICs | 0 | 0.00% |

Abbreviations: UK = United Kingdom, HU = Hungary, NUH = Nottingham University Hospitals NHS Trust, LTHT = Leeds Teaching Hospital NHS Trust, ULH = United Lincolnshire Hospitals NHS Trust, KAP= Budapest, BUS = Szűrőbusz, SZO = Szolnok, KKM = Kecskemét, SZE = Szekszárd, ICs = interval cancers

**Table S2:** Cancer detection performance per clinical site

| NUH (UK) |  |  |  |  |  |  |  |  |
| --- | --- | --- | --- | --- | --- | --- | --- | --- |
|  | Historical HDR |  |  | Simulated AI-IR / AI-SR |  |  | Test AI vs HDR |  |
| Performance metric | Value | LBCI | UBCI | Value | LBCI | UBCI | Ratio | Difference |
| Recall rate | 2.84 | 2.71 | 2.96 | 2.88 | 2.76 | 3.00 | Non-inferior | Non-inferior |
| CDR | 9.05 | 8.34 | 9.77 | 8.85 | 8.14 | 9.57 | Non-inferior | Non-inferior |
| Sensitivity | 85.83 | 82.90 | 88.45 | 83.92 | 81.03 | 86.53 | Non-inferior | Non-inferior |
| Specificity | 97.87 | 97.67 | 98.06 | 97.91 | 97.72 | 98.11 | Non-inferior | Non-inferior |
| PPV | 31.91 | 29.78 | 34.09 | 30.75 | 28.63 | 32.87 | Non-inferior | Non-inferior |
| LTHT (UK) |  |  |  |  |  |  |  |  |
|  | Historical HDR |  |  | Simulated AI-IR / AI-SR |  |  | Test AI vs HDR |  |
| Performance metric | Value | LBCI | UBCI | Value | LBCI | UBCI | Ratio | Difference |
| Recall rate | 5.09 | 4.93 | 5.27 | 5.06 | 4.89 | 5.23 | Non-inferior | Non-inferior |
| CDR | 8.34 | 7.67 | 9.12 | 8.13 | 7.45 | 8.88 | Non-inferior | Non-inferior |
| Sensitivity | 87.30 | 84.81 | 89.87 | 85.05 | 82.39 | 87.69 | Non-inferior | Non-inferior |
| Specificity | 95.92 | 95.68 | 96.18 | 96.01 | 95.76 | 96.27 | Non-inferior | Non-inferior |
| PPV | 16.38 | 15.13 | 17.76 | 16.07 | 14.85 | 17.45 | Non-inferior | Non-inferior |
| ULH (UK) |  |  |  |  |  |  |  |  |
|  | Historical HDR |  |  | Simulated AI-IR / AI-SR |  |  | Test AI vs HDR |  |
| Performance metric | Value | LBCI | UBCI | Value | LBCI | UBCI | Ratio | Difference |
| Recall rate | 3.63 | 3.47 | 3.78 | 3.56 | 3.42 | 3.71 | Superior | Superior |
| CDR | 9.44 | 8.65 | 10.20 | 9.19 | 8.41 | 9.93 | Non-inferior | Non-inferior |
| Sensitivity | 85.65 | 83.11 | 88.27 | 83.36 | 80.70 | 86.13 | Non-inferior | Non-inferior |
| Specificity | 97.37 | 97.05 | 97.68 | 97.49 | 97.17 | 97.78 | Non-inferior | Non-inferior |
| PPV | 26.00 | 24.12 | 27.96 | 25.82 | 23.97 | 27.68 | Non-inferior | Non-inferior |
| KAP (HU) |  |  |  |  |  |  |  |  |
|  | Historical HDR |  |  | Simulated AI-IR / AI-SR |  |  | Test AI vs HDR |  |
| Performance metric | Value | LBCI | UBCI | Value | LBCI | UBCI | Ratio | Difference |
| Recall rate | 11.43 | 11.03 | 11.82 | 9.95 | 9.56 | 10.31 | Superior | Superior |
| CDR | 7.22 | 6.24 | 8.23 | 7.11 | 6.16 | 8.12 | Non-inferior | Non-inferior |
| Sensitivity | 82.92 | 78.11 | 87.80 | 81.67 | 76.99 | 86.49 | Non-inferior | Non-inferior |
| Specificity | 94.11 | 93.62 | 94.61 | 95.37 | 94.93 | 95.80 | Superior | Superior |
| PPV | 6.31 | 5.46 | 7.17 | 7.15 | 6.20 | 8.11 | Superior | Superior |
| BUS (HU) |  |  |  |  |  |  |  |  |
|  | Historical HDR |  |  | Simulated AI-IR / AI-SR |  |  | Test AI vs HDR |  |
| Performance metric | Value | LBCI | UBCI | Value | LBCI | UBCI | Ratio | Difference |
| Recall rate | 6.25 | 5.93 | 6.57 | 5.37 | 5.08 | 5.68 | Superior | Superior |
| CDR | 4.56 | 3.68 | 5.40 | 4.52 | 3.68 | 5.31 | Non-inferior | Non-inferior |
| Sensitivity | 87.20 | 81.02 | 92.62 | 86.40 | 80.00 | 92.24 | Non-inferior | Non-inferior |
| Specificity | 96.06 | 95.19 | 96.84 | 96.58 | 95.85 | 97.34 | Superior | Superior |
| PPV | 7.30 | 5.98 | 8.56 | 8.41 | 6.88 | 9.82 | Superior | Superior |
| **SZO (HU)** |  |  |  |  |  |  |  |  |
|  | **Historical HDR** |  |  | **Simulated AI-IR / AI-SR** |  |  | **Test AI vs HDR** |  |
| **Performance metric** | **Value** | **LBCI** | **UBCI** | **Value** | **LBCI** | **UBCI** | **Ratio** | **Difference** |
| Recall rate | 18.21 | 17.68 | 18.75 | 16.50 | 15.98 | 17.02 | Superior | Superior |
| CDR | 13.65 | 11.99 | 15.21 | 13.50 | 11.83 | 15.05 | Non-inferior | Non-inferior |
| Sensitivity | 92.93 | 89.90 | 95.82 | 91.87 | 88.64 | 94.91 | Non-inferior | Non-inferior |
| Specificity | 94.14 | 93.23 | 95.04 | 95.38 | 94.56 | 96.18 | Superior | Superior |
| PPV | 7.50 | 6.61 | 8.30 | 8.18 | 7.26 | 9.08 | Superior | Superior |
| **KKM (HU)** |  |  |  |  |  |  |  |  |
|  | **Historical HDR** |  |  | **Simulated AI-IR / AI-SR** |  |  | **Test AI vs HDR** |  |
| **Performance metric** | **Value** | **LBCI** | **UBCI** | **Value** | **LBCI** | **UBCI** | **Ratio** | **Difference** |
| Recall rate | 14.19 | 13.54 | 14.85 | 12.55 | 11.92 | 13.16 | Superior | Superior |
| CDR | 7.42 | 5.89 | 8.96 | 7.34 | 5.80 | 8.96 | Non-inferior | Non-inferior |
| Sensitivity | 89.69 | 82.88 | 95.00 | 88.66 | 81.44 | 94.32 | Non-inferior | Non-inferior |
| Specificity | Insufficient follow-up data for reliably reporting specificity |  |  |  |  |  |  |  |
| PPV | 5.23 | 4.15 | 6.36 | 5.85 | 4.62 | 7.03 | Superior | Superior |
| **SZE (HU)** |  |  |  |  |  |  |  |  |
|  | **Historical HDR** |  |  | **Simulated AI-IR / AI-SR** |  |  | **Test AI vs HDR** |  |
| **Performance metric** | **Value** | **LBCI** | **UBCI** | **Value** | **LBCI** | **UBCI** | **Ratio** | **Difference** |
| Recall rate | 8.51 | 7.68 | 9.36 | 6.80 | 6.02 | 7.61 | Superior | Superior |
| CDR | 6.78 | 4.27 | 9.54 | 6.78 | 4.27 | 9.54 | Non-inferior | Non-inferior |
| Sensitivity | 100.00 | 100.00 | 100.00 | 100.00 | 100.00 | 100.00 | Non-inferior | Non-inferior |
| Specificity | Insufficient follow-up data for reliably reporting specificity |  |  |  |  |  |  |  |
| PPV | 7.96 | 5.09 | 11.08 | 9.96 | 6.46 | 13.86 | Superior | Superior |

Abbreviations: UK = United Kingdom, HU = Hungary, NUH = Nottingham University Hospitals NHS Trust, LTHT = Leeds Teaching Hospital NHS Trust, ULH = United Lincolnshire Hospitals NHS Trust, KAP= Budapest, BUS = Szűrőbusz, SZO = Szolnok, KKM = Kecskemét, SZE = Szekszárd, HDR = human double reading, AI-IR = AI independent reader, AI-SR = AI supporting reader, LBCI = lower bound 95% confidence interval, UBCI = upper bound 95% confidence interval, CDR = cancer detection rate (per 1000 screens), PPV = positive predictive value

